# Evaluation of current practice of antimicrobial use and clinical outcome in the management of children with pneumonia admitted to Jimma Medical Center, Southwest Ethiopia: A prospective observational study

**DOI:** 10.1101/2024.10.26.24316179

**Authors:** Abduba Wariyo Guyo, Berhanu Teshome Derese

**Affiliations:** Department of Pharmacy, Institute of Health, Bule Hora University, Bule Hora, Ethiopia; Department of Pharmacy, College of Medicine and Health Science, Salale University, Salale, Ethiopia

**Keywords:** Antimicrobial use, Current practice, Jimma Medical Center, Pneumonia

## Abstract

**Background:** Antimicrobial resistance is a global crisis that threatens to reverse a century of medical progress; threatening the effective prevention and treatment of common infectious diseases.

**Objectives:** To evaluate the current practices of antimicrobial utilization and clinical outcomes of children with pneumonia admitted to Jimma Medical Center, Ethiopia.

**Method:** A prospective observational study design was conducted on children admitted to the pediatric wards of Jimma Medical Center. The study was conducted from February 03, 2022, to June 03, 2022, and patients aged < 18 years and diagnosed with pneumonia were included. A chart review supplemented by a self-administered questionnaire was used to collect data. Descriptive statistics and binary logistic regressions were performed for data analyses.

**Results:** Among the total of 146 patients, 61.6% were male, and the mean age was 40.95 (+47.61) months. Microbiologically and radiologically examined patients were 47(32.19%) and 64 (43.8%), respectively. All the treatment approaches were found to be initiated empirically. Ceftriaxone was the most commonly prescribed antimicrobial. In-hospital mortality was 7.5%. Pre-admission antimicrobial use (AOR =3.87; 95% CI:1.34-11.16; P=0.012), antimicrobial change (AOR = 3.74; 95% CI: 1.522-9.22; P=0.004), and hospital stay (> 10 days) (AOR = 6.00; 95% CI: 2.53-14.22; P=0.029) were all independent predictors of poor clinical outcome.

**Conclusion:** An empirically initiated antimicrobial was completed without sufficient evidence of indication, such as microbiological and radiographic findings. More than one-fourth of the patients treated for pneumonia experienced poor outcomes, implicating the need for more attention during treatment.

## 1. Introduction

The discovery of antimicrobial agents is one of human history’s most important medical advances to reduce disease and save billions of lives [1][2]. However, the successful use of antimicrobial agents is compromised by misuse [3][4][5], and the development of resistance in the past few years [5]. Antimicrobial resistance is a global crisis that threatens to reverse a century of medical progress; threatening the effective prevention and treatment of common infections, leading to increased morbidity and mortality, and associated economic costs due to the health care burden [6][7]. Reports indicate that there will be about 10 million AMR-related deaths every year until 2050, with the majority being in Africa and Asia [8].

Inappropriate antimicrobial use is a major driver of antimicrobial resistance (AMR). Earlier Studies conducted in pediatric wards in Ethiopian hospitals have shown inappropriate use of antimicrobials [9][10][11]. In these studies, one of the main reasons for inappropriate antimicrobial use was the management of pneumonia. Pneumonia is the leading cause of morbidity and mortality in Ethiopia, especially in the pediatric population, and its main factors lead to hospitalization and intensive use of antimicrobials [12].

In addition to the AMR, Inappropriate and indiscriminate use of antimicrobials leads to undesirable consequences, including severe illnesses, prolonged hospitalization, increased morbidity and mortality, increased Clostridium difficile and other hospital-acquired infections (HAIs), and increased healthcare expenses [6][13][14].

The Ministry of Health and its subordinates, in particular, the Ethiopian Food and Drug Administration (EFDA), have worked together to tackle antimicrobial resistance by developing national drug policies [16], developing standard treatment guidelines for the different healthcare establishments at different times [17], drug formulary, good prescription and dispensing manuals [18] and different practical guides since the issuance of declaration 661 2009. All of these efforts are being made to tackle AMR by promoting rational antimicrobial use and protecting citizens [19]. However, the effective implementation of standards and most treatment guidelines remains unexplored. Therefore, this study attempted to evaluate antimicrobial use in current practice and clinical outcomes in the management of childhood pneumonia. Hence, this helps provide specific information for designing an appropriate intervention strategy that optimizes antimicrobial use and improves economic and patient outcomes. Ultimately, the findings may call for policymakers for effective drug regulation in Ethiopia.

## 2. Material and Methods

### 2.1 Ethical Statement

The proposal including Amharic and Oromic written assent, which was attached as an annex, was submitted to the School of Pharmacy, Jimma University Institutional Review Board for review and approval. The study was conducted after securing the letter of ethical approval (Ref. No. JUIRB016/14). A formal letter was written to Jimma Medical Center (Ref. No.4/7/n7/133/2014/) and, the hospital medical director and head of the department of pediatric ward were informed about the purpose of the study to get permission and cooperation. Written informed from the patient’s Parent or guardian was obtained after providing information regarding the purpose/nature of the study. Parents or guardians were told the reasons for their child being selected to be included in the study and assured that declining participation would not have any influence on the right of their child to get treatment. The parent or guardian was also told about the right to withdraw from the study at any time. The parent or guardian was assured of confidentiality (privacy and anonymity) of the information obtained during the study.

### 2.2 Study Setting and Period

The study was conducted from February 03, 2022, to June 03, 2022, in the pediatric ward of Jimma Medical Center (JMC), a teaching and referral hospital located in Jimma town, Oromia Region in southwestern Ethiopia, 352 km from the national capital, Addis Ababa. JMC provides services for approximately 15,000 inpatients, 160,000 outpatient attendants, and 11,000 emergency cases per year. The hospital serves more than 20 million people coming to the hospital from the catchment area. It has different wards. The pediatric ward is among the wards that have different units such as level I, level II, neonatal unit, intensive care unit, and oncology unit.

### 2.3 Study Design

A hospital-based prospective observational study was conducted on children with pneumonia admitted to the pediatric ward of Jimma Medical Center.

### 2.4 Participant Eligibility and Inclusion

All pediatric patients admitted to the pediatric ward of the Jimma Medical Center with a diagnosis of pneumonia during the study period were screened for eligibility criteria. Patients aged less than or equal to 18 years admitted to the pediatric wards of Jimma Medical Center with a suspected or proven pneumonia diagnosis by physician decision on duty during the study period and Patients whose parents gave assent were included in the study. Patients who died before they started antimicrobial, transferred to other facilities and patient lost follow-up were excluded.

### 2.5 Sample Size and Sampling Technique

No sampling technique was applied; instead, all pediatric patients diagnosed with pneumonia who fulfilled illegibility criteria during the study period were recruited in the study. Accordingly, 146 patients were followed and completed data collected.

### 2.6 Data Collection Instrument and Quality Assurance

The data collection instrument (data abstraction format and self-administered questionnaire) was developed by reviewing the literature on antimicrobial use, antimicrobial resistance, and antimicrobial stewardship program guidelines. Training was given to data collectors on the data collection procedure and research objectives. The principal investigator was regularly checking the consistency and completeness of the data and random checks were also being performed.

### 2.7 Data Processing and Analysis

Data was compiled, cleared, coded, and verified for completeness and accuracy before being included in Epi Data Manager version 4.6.2. Double-entry verification was performed and the data was analyzed using statistical software, SPSS version 25.0. Descriptive statistics (mean, frequency, and cross-tabulation) were used to describe the practice of antimicrobial use. The bivariate logistic regression was performed for clinical outcome and variables with p-values less than 0.25 with outcomes (poor outcome or good outcome) were taken into account for further analysis. Then, the independent predictors were identified by multivariate logistic regression analysis. All statistical tests were detailed; a P value of less than or equal to 0.05 was considered statistically significant.

## 3. Results

### 3.1 Socio-demographic Data

A total of 161 children admitted to the pediatric ward of JMC were diagnosed with pneumonia and began antimicrobial treatment for pneumonia during the study period. However, data for 15 patients was not complete as a result of discharges against medical advice and some lost follow-up (Fig 1).

**Fig 1.**
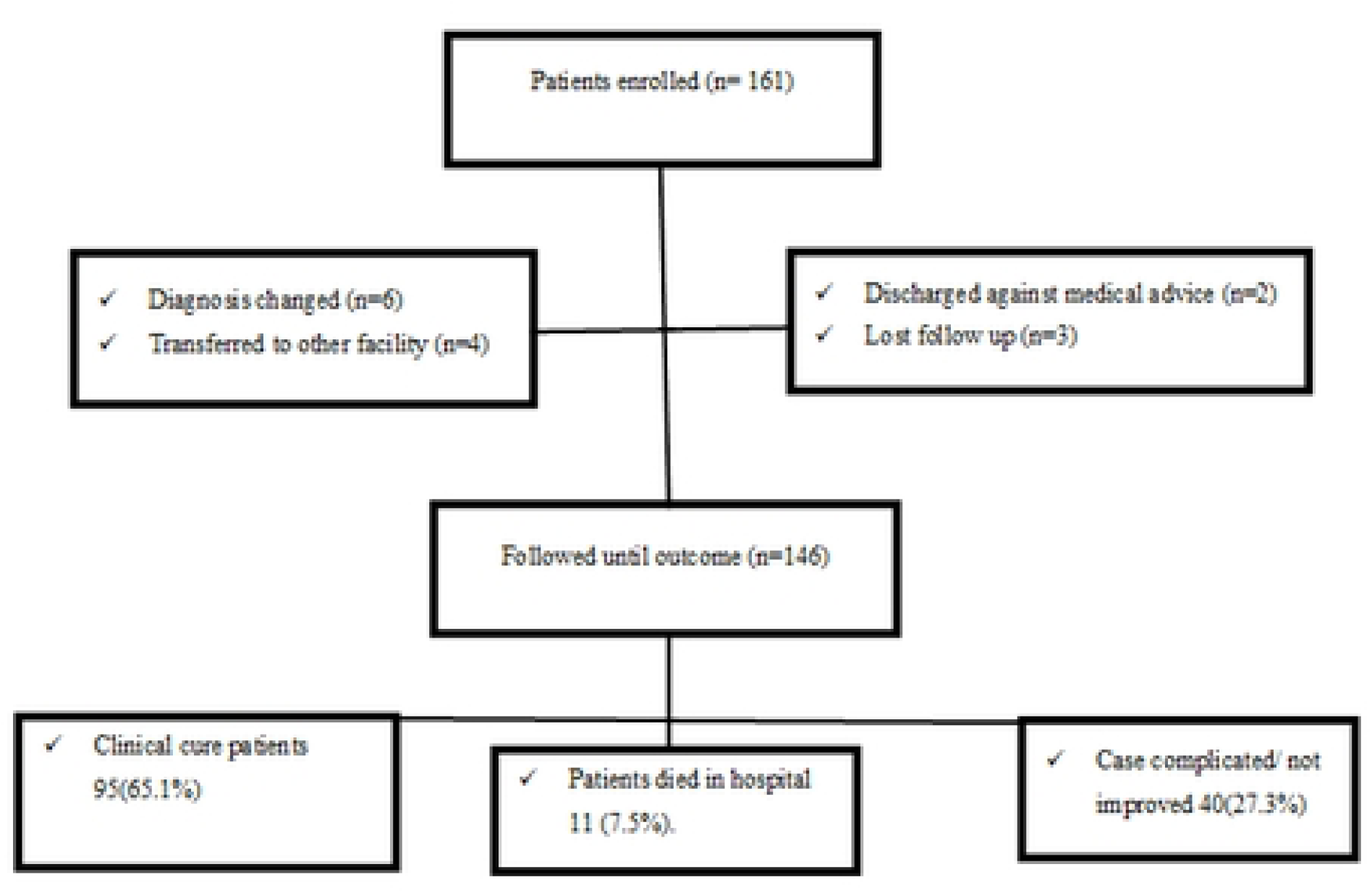
Flow diagram showing enrollment of children with pneumonia at Jimma Medical Center, Jimma, Ethiopia, February 03 to June 03, 2022

According to this study, the mean age of participants was 40.95 (+47.61) months, with more than half (61.6%) of male children. most of the participants, 65 (44.5%), were in the age range less than 11 months. The socio-demographic data is in Table 1.

**Table 1.** Socio-demographic characteristics of children with pneumonia at Jimma Medical Center, Jimma, Ethiopia, February 03 to June 03, 2022.

| Variable | Value | Frequency | % |
| --- | --- | --- | --- |
| Sex | Male | 90 | 61.6 |
|  | Female | 56 | 38.4 |
| Age category (in months) | < =11 | 65 | 44.5 |
|  | 12-35 | 22 | 15.1 |
|  | 36-71 | 31 | 21.2 |
|  | >=72 | 28 | 19.2 |
| Age, months | (Mean ± SD) | (40.95 ± 47.61) | |
| Weight, Kg | 1-5.9 | 29 | 19.9 |
|  | 6-10.9 | 59 | 40.4 |
|  | 11-15.9 | 33 | 22.6 |
|  | >=16 | 25 | 17.1 |
| Residence | Rural | 129 | 88.4 |
|  | Urban | 17 | 11.6 |
| Type of admission | Direct Admission | 11 | 7.5 |
|  | Referred from gov't institution | 120 | 82.2 |
|  | Referred from private institute | 15 | 10.3 |

### 3.2 Clinical Characteristics

More than three-quarters (76%) of patients had comorbidities alongside pneumonia. The two most common reasons for admissions were malnutrition + pneumonia and pneumonia with frequencies of 37(25.3%) and 35(23.9%). Malnutrition and cardiovascular disease were the most common comorbidities in the study population, accounting for 52 (35.6%) and 30(20.5%), respectively (Table 2).

**Table 2.**
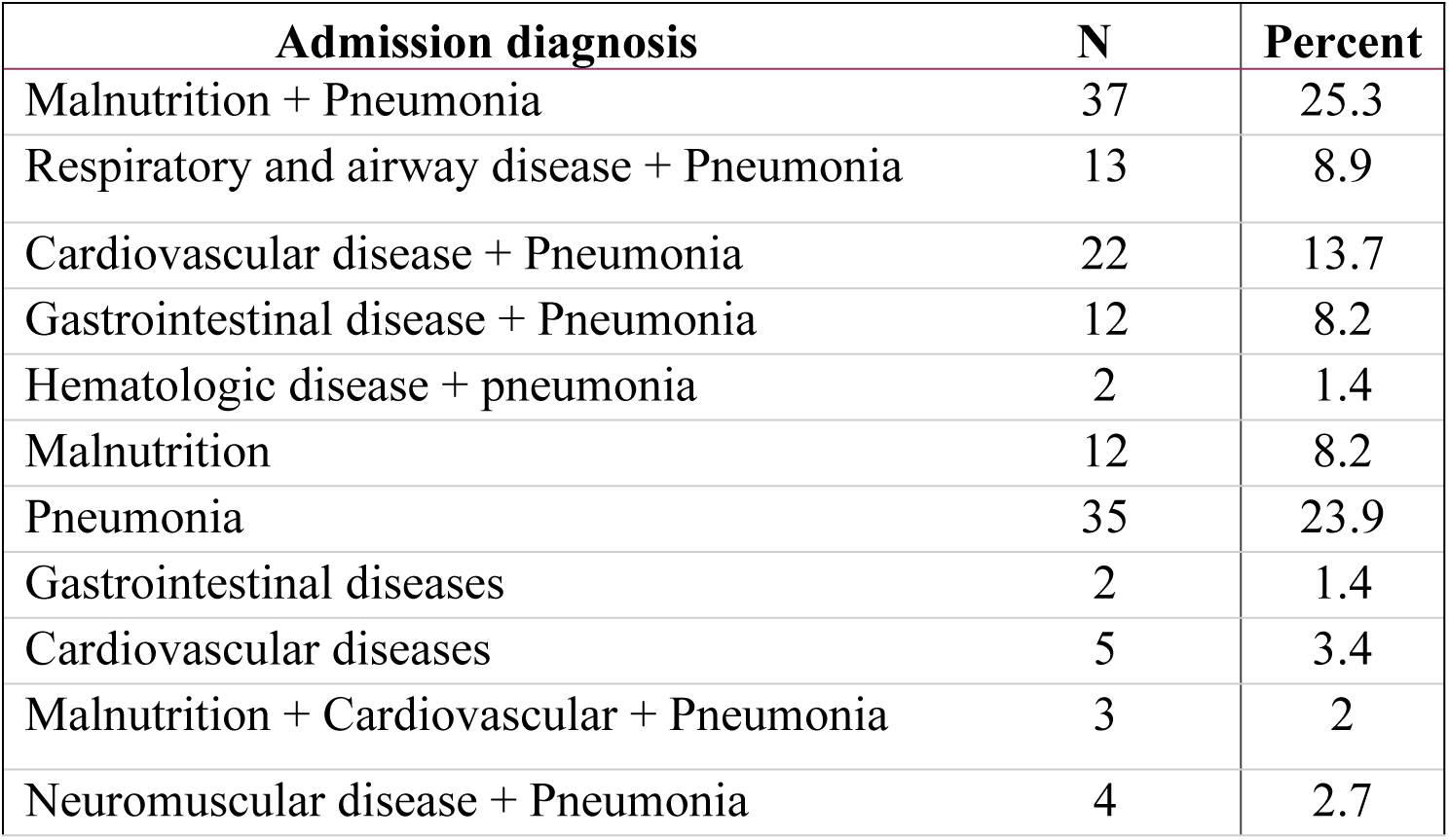
Pneumonia patients’ admission diagnosis and their frequency at Jimma Medical Center, Jimma, Ethiopia, from February 03 to June 03, 2022.

The majority of participants (89, 61%) of them were having Pre-admit antimicrobial use. More than three-quarters of patients (120, 82.2%), were diagnosed with community-acquired pneumonia and the majority (121,82.9%) of patients had severe pneumonia. Baseline clinical characteristics in Table 3.

**Table 3.**
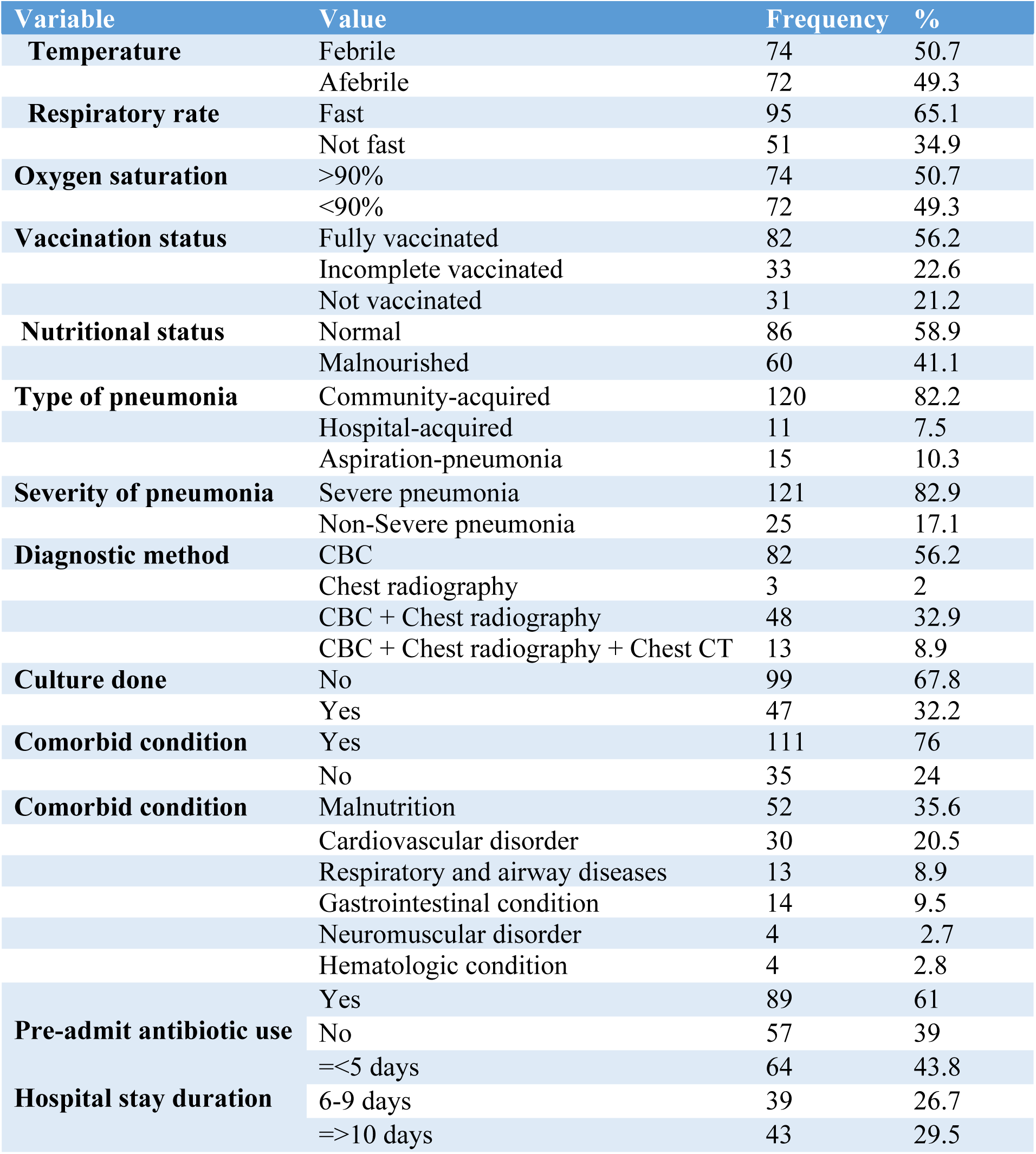
Baseline clinical characteristics of children with pneumonia at Jimma Medical Center, Ethiopia, from February 03 to June 03, 2022.

### 3.3 Practice of microbiology studies

Among the 146 children diagnosed with pneumonia and hospitalized and given antimicrobials, microbiologic testing was performed only for 47(32.19%) children. Of the cultured microbiology samples, 16 (34.0%) were collected before the initiation of empiric antimicrobial therapy. The most common samples taken were pleural fluid, 24(16.4%), followed by blood, 11 (7.5%). The yield of microbiological culture was low in this study. Only thirteen bacterial species, which were designated as probable pathogens, were isolated from 28.2% (13/47) of the cultures (fig 2).

**Figure 2.**
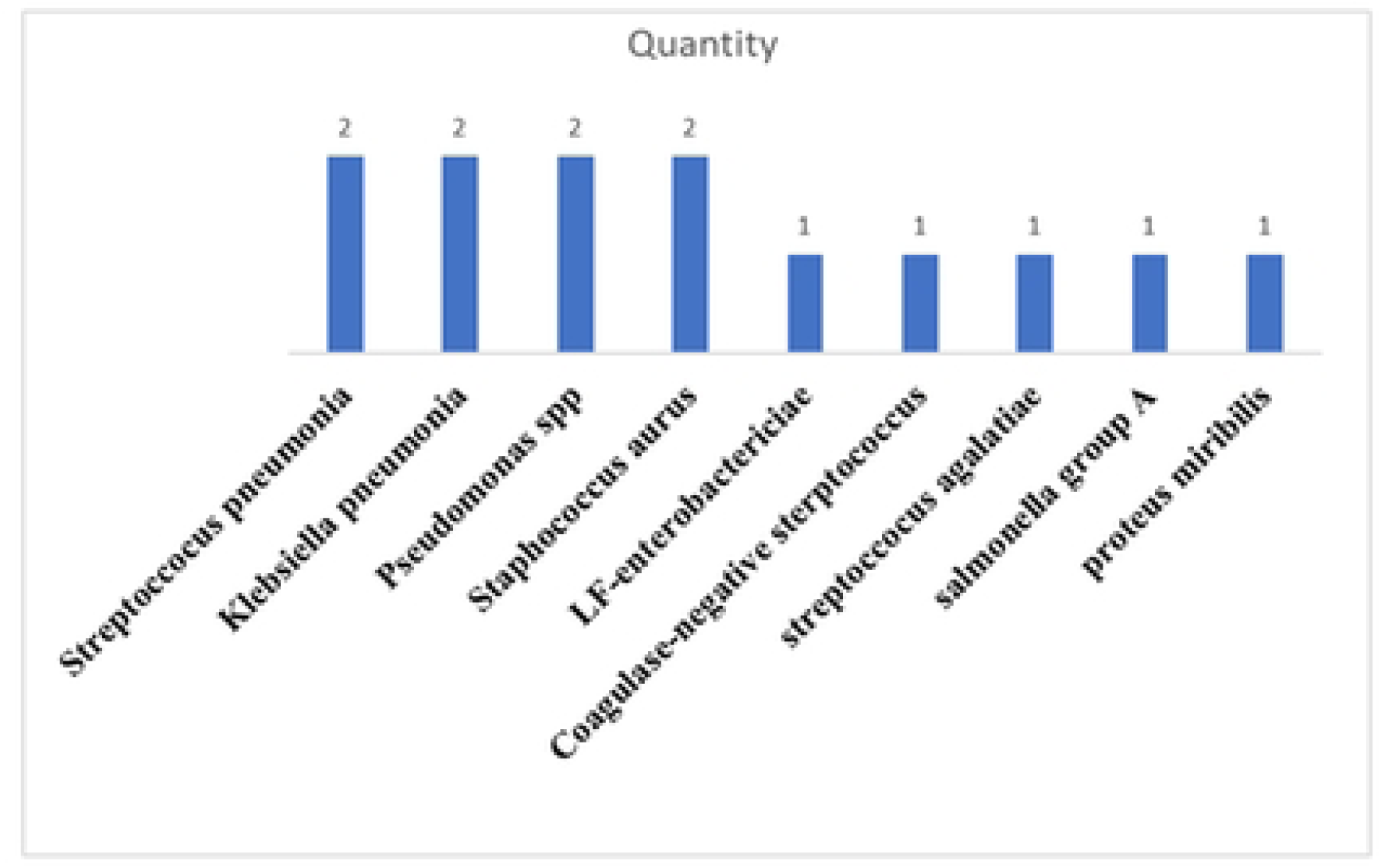
Identified organisms during the management of children with pneumonia at Jimma Medical Center, Jimma, Ethiopia, February 03 to June 03, 2022.

Fifteen treating physicians who do not order microbiologic studies wrote down their reasons why they didn’t send them, and more than half said that they did not order them because " it is usual practice to treat childhood pneumonia based on clinical presentation" (Table 4).

**Table 4.** Treating physicians’ reasons for not considering microbiological studies for the management of pneumonia at Jimma Medical Center, Jimma, Ethiopia, February 3 to June 3, 2022.

| Reason given by physicians | N (%) |
| --- | --- |
| The usual practice is to treat children pneumonia is based only on clinical (Signs and symptoms) | 9(60.1) |
| Patient already started antimicrobial | 2(13.3) |
| Culture result always negative and only radiologic information is sufficient to treat pneumonia in pediatrics | 2(13.3) |
| It takes time waiting lab. Result | 2(13.3) |

### 3.4 Antimicrobial prescribing pattern and susceptibility

As shown in Figure 3, it can be observed that about 7 types of antimicrobials were used for the treatment of different types of pneumonia. Out of 146 patients, 95(65.1%) of the patients received ceftriaxone, and it was the most commonly prescribed empiric antimicrobial agent for pneumonia. Regarding polypharmacy, the average number of antimicrobial prescriptions per patient was 1.68, which means that almost all of the patients were exposed to at least 2 antimicrobial agents regardless of the type of pneumonia.

**Figure 3.**
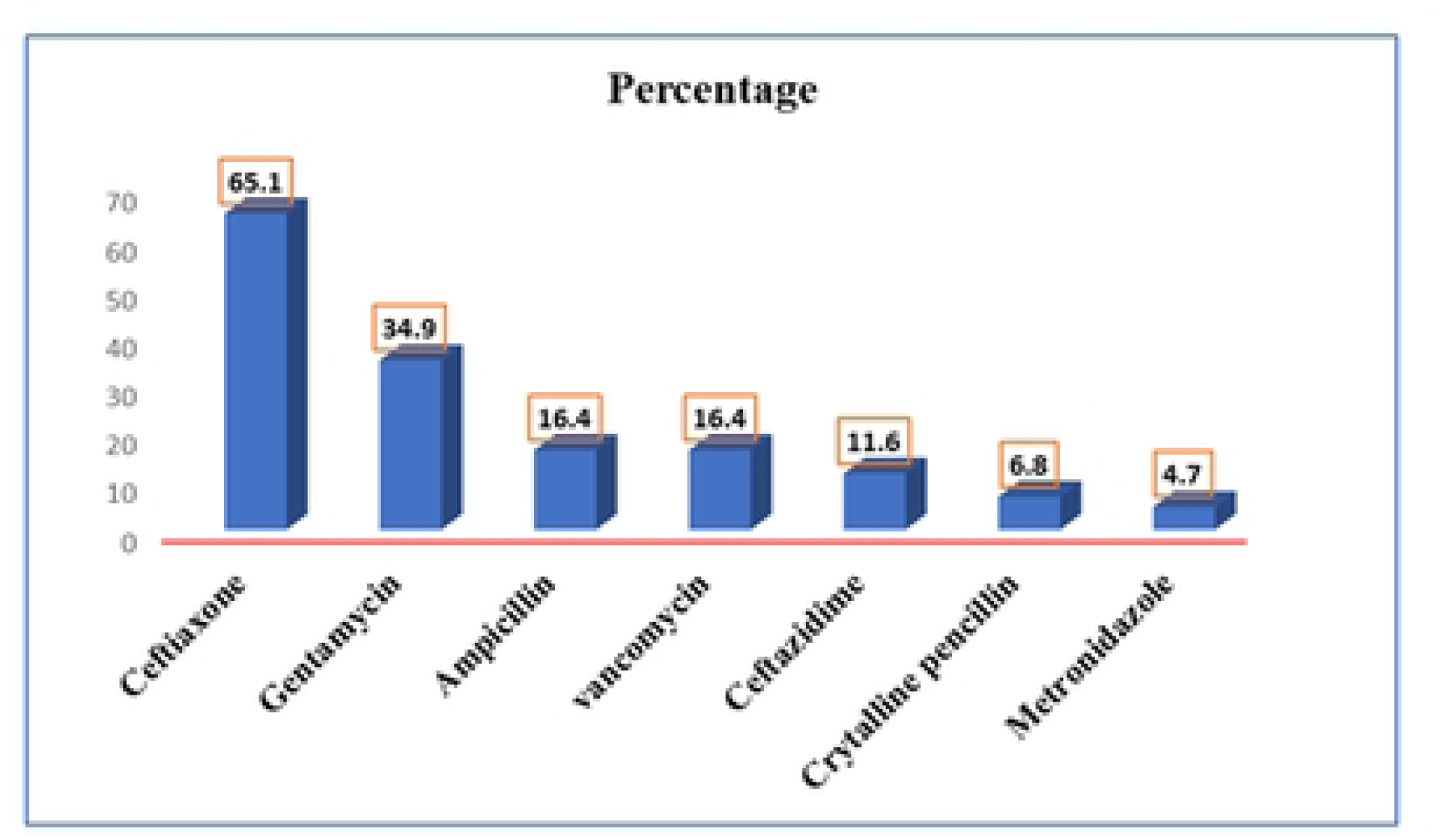
The most commonly used empiric antimicrobials (percent) for the management of children with pneumonia at Jimma Medical Center, Jimma, Ethiopia, February 03 to June 03, 2022

In this study, all the treatment approaches were found to be initiated empirical. All patients diagnosed with pneumonia got either an antimicrobial regimen alone or in a combination of two or more antimicrobials for all types of pneumonia. Among 146 patients diagnosed with pneumonia, it can be seen that 67 patients received single antimicrobial therapy and other patients received combinations of two or three antimicrobial therapies as initial antimicrobial regimens. Ceftriaxone was the most widely used monotherapy 58 (39%) and in combination with another antimicrobial 37 (25.3%). The type of pneumonia, in terms of antimicrobial selection, varied among patients with similar diagnoses (table 5).

**Table 5.** Types of pneumonia and the initial antimicrobial regimens that patients received at Jimma Medical Center, Jimma, Ethiopia, February 03 to June 03, 2022.

| Antimicrobial used as first-line regimen | CAP | HAP | AP | Total |
| --- | --- | --- | --- | --- |
| Ampicillin + gentamycin | 24 | - | - | 24 |
| Ceftriaxone | 47 | 5 | 5 | 57 |
| Ceftriaxone + Azithromycin | 2 | 1 | 1 | 4 |
| Ceftriaxone + Gentamycin | 20 | 5 | 2 | 27 |
| Ceftriaxone + Vancomycin | 4 | - | - | 4 |
| Ceftazidime + Vancomycin | 9 | - | 4 | 13 |
| C. penicillin | 10 | - | - | 10 |
| Ceftriaxone + Vancomycin + Metronidazole | 2 | - | 1 | 3 |
| Ceftazidime + Vancomycin + Metronidazole | 2 | - | 2 | 4 |
| Total | 120 | 11 | 15 | 146 |

It was assessed whether there was a change in the initial antimicrobial regimen or not during treatment, and it was found that there were 64(43.8%) first-time changes, and the most common reason associated with the changes was a poor response to the initial antimicrobials (fig 4).

**Figure 4.**
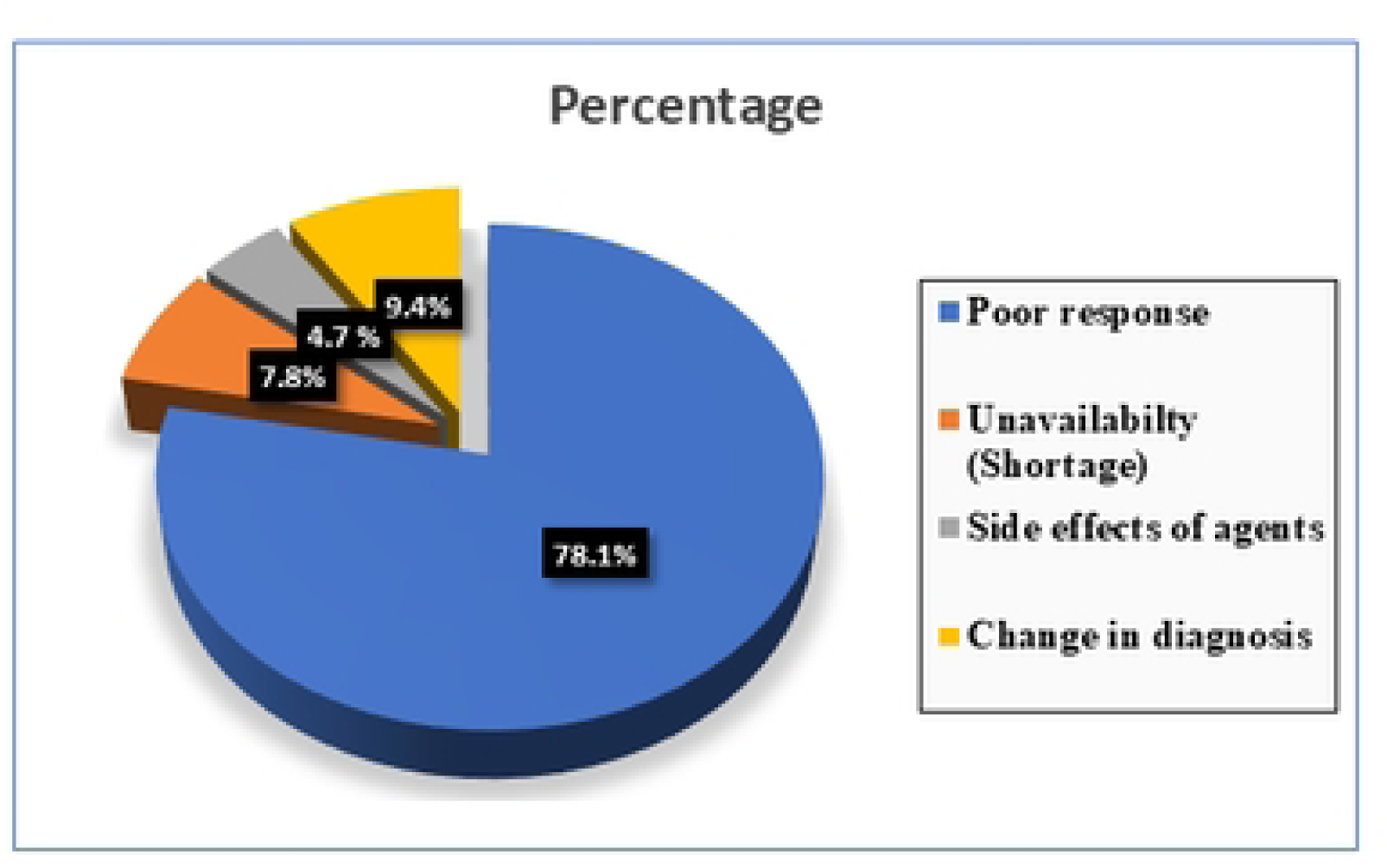
Reasons for change in initial antimicrobial regimen for the management of children with pneumonia at Jimma Medical Center, Jimma, Ethiopia, February 03 to June 03, 2022.

Out of a total of 47 microbiology tests done, only 13(28.2%) showed bacterial growth. The majority of the detected organisms were found to be resistant to the commonly prescribed Beta-lactam antimicrobials for the treatment of pneumonia based on the susceptibility data collected from the 13 culture-positive results (table 6).

**Table 6.** The list of identified pathogens and their susceptibility patterns to selected antimicrobial in the management of children with pneumonia at Jimma Medical Center, Ethiopia, from February 03 to June 03, 2022.

|  | Coagulase-negative<br>streptococcus<br>(n=1) | Streptococcus<br>agalatae (n=1) | K. pneumonia<br>(n=2) | Pseudomonas<br>Spp. (n=2) | S. aureus<br>(n=2) | LF-Entero<br>bacteri<br>aceae<br>(n= 1) | Salmonella<br>group A<br>(N=1) | Proteus<br>mirabilis<br>(n=1) | Streptococcus<br>Pneumonia<br>(n=2) |
| --- | --- | --- | --- | --- | --- | --- | --- | --- | --- |
| Amoxicillin | - | - | R | R | - | - | R | R | R |
| Ampicillin | - | S | R | R | - | - | R | R | R |
| C. penicillin | - | S | - | - | - | - | - | - | - |
| Gentamycin | - | - | R | S | R | S | S | - | - |
| Azithromycin | - | - | - | - | R | - | - | - | - |
| Ceftriaxone | - | - | R | R | R | - | R | - | R |
| ceftazidime | - | - | - | - | - | - | R | R | R |
| vancomycin | R | - | - | - | S | - | - | - | S |
| CAF | S | - | S | S | S | - | S | S | S |
| Oxacillin | R | - | - | - | S | - | - | - | R |
| Erythromycin | - | R | - | - | R | - | - | - | R |
| Meropenem | - | - | S | - | S | S | - | - | - |
| TTC | - | R | - | R | R | S | - | - | - |
| Cefazoline |  |  | R |  | R |  |  |  | R |
| Cotrimoxazole | - | - | - | R | S | S | S | - | - |
| P/T | - | - | S | R | - | S | - | S | S |
| Cefotaxime | - | - | - |  | - | - | S | - | - |
| Imipenem | - | - | S | S | - | - | S | - | S |

P/T= Piperacillin/tazobactam, TTC= Tetracycline, CAF= Chloramphenicol, R=Resistant, S=Susceptible

The appropriateness or adequacy of the initial antimicrobial regimens is described in light of the identified microorganisms (table 7).

**Table 7.** Organisms isolated, susceptibility data, and initial antimicrobial therapy to demonstrate adequacy and appropriateness at Jimma Medical Center, Ethiopia, from February 03 to June 03, 2022.

| Identified organism | Susceptible to | Initial antimicrobial regimen given | Appropriateness/ Adequacy (Yes/No) |
| --- | --- | --- | --- |
| Streptococcus pneumonia | Vancomycin, Imipenem | Ceftriaxone | No |
|  | piperacillin/tazobactam, chloramphenicol | Ampicillin and Gentamycin | No |
| Klebsiella pneumonia | Meropenem, Chloramphenicol | Ceftriaxone | No |
|  | Piperacillin/tazobactam, Imipenem, | Ceftriaxone | No |
| Pseudomonas spp. | Ceftazidime, Cefuroxime, Imipenem, P/T, CAF, Gentamycin | Ceftriaxone and Gentamycin | No/Yes |
|  | Meropenem, Chloramphenicol, P/T | Ceftazidime and Vancomycin | - |
| Staphylococcus aureus | Oxacillin, Gentamycin, CAF, TTC, ciprofloxacin, Clindamycin, Cotrimoxazole, Erythromycin | Ampicillin and Gentamycin | Yes |
|  | Oxacillin, Penicillin, Gentamycin, Ciprofloxacin | Crystalline penicillin | Yes |
| LF-Enterobacteriaceae | Cotrimoxazole, P/T, TTC, Meropenem | Ceftriaxone | No |
| Coagulase-negative staphylococcus | Chloramphenicol | Ceftriaxone and vancomycin | -/No |
| Streptococcus agalatae | Ampicillin, penicillin | Ampicillin and Gentamycin | Yes/- |
| Salmonella group A | Cotrimoxazole, Gentamycin, Cefotaxime, Imipenem, CAF | Ceftriaxone and Gentamycin | No/Yes |
| proteus miribilis | P/T, CAF | Ceftriaxone | No |

P/T= Piperacillin/tazobactam, CAF= Chloramphenicol, TTC= Tetracycline

The most common prescribing physicians were residents (12, 63.2%), medical interns (5, 26.3%), and specialists (2, 10.5%). The guidelines/protocol used for prescribing antimicrobials for the management of children with pneumonia were treating physician asked and the most common guideline to prescribe antimicrobials was reference eBooks for 13(68.4%), international guidelines (IDSA/BTS) 4 (21.1%), and WHO guideline 2(10.5%). The prescribing physicians were asked whether they consulted a clinical pharmacist in the selection of appropriate antimicrobials and their response was no. The reasons for not consulting were also asked, and all of them wrote down their reasons for not consulting a clinical pharmacist in the management of pneumonia, and the commonest reasons were summarized as follows: The most common reasons were: unavailability of clinical pharmacists in the wards (12, 63.1%), no assigned clinical pharmacist (5, 26.3%), and no need to consult a clinical pharmacist (2, 10.5%).

### 3.5 Switching from Intravenous to Oral Therapy

Patients’ eligibility for IV to PO conversion was assessed, and it was found that 32 patients were eligible for conversion; Only 3 were transferred, and the rest continued to receive intravenous IV antimicrobial. All of the physicians treating the patients during the study period were asked about the practice of IV to PO conversion, and the response was no. Almost all of them wrote down their reasons for not practicing IV to PO conversion in the datasheet, and the most typical reason was "conversion only made on at discharge" 7(36.8%) (table 8).

**Table 8.** Reasons for not practicing IV to PO conversion in the management of children with pneumonia at Jimma Medical Center, Jimma, Ethiopia, February 03 to June 03, 2022.

| Reason given by physician | N (%) |
| --- | --- |
| Conversion only made at discharge | 7(36.8) |
| The usual practice is to treat admitted children with IV antimicrobial | 5(26.3) |
| It is better to complete it with an intravenous | 4(21.1) |
| Other | 3(15.7) |

### 3.6 Clinical outcome

All the patients were followed starting from the initiation of antimicrobial therapy till a clinical outcome was achieved and antimicrobial therapy was discontinued. The patient’s clinical outcome was observed immediately after the end of treatment, and the physicians who followed the patient were asked about their clinical status following antimicrobial treatment. Accordingly, the observed outcomes were: clinical cure 95(65.1%) and complications 40(27.3%). The total in hospital mortality was 11 (7.5%). The results, especially the clinical cured, have not been confirmed by an independent physician and may be biased towards respondents (table 9).

**Table 9.** The type of pneumonia and clinical outcomes of children with pneumonia at Jimma Medical Center from February 03 to June 03, 2022.

| Type of pneumonia | Outcome status |  |  |
| --- | --- | --- | --- |
|  | Good outcome | Poor outcome | Total |
| Community-acquired | 82 | 36 | 118 |
| Hospital-acquired | 6 | 5 | 11 |
| Aspiration-pneumonia | 7 | 10 | 17 |
| Total | 95 | 51 | 146 |

### 3.7 Predictors of Poor Outcome in Children with Pneumonia

Initially, the clinical outcome was categorized into good outcomes (clinical cured and improved patients) and poor outcomes (death and complicated patients) because a logistic regression model requires the dependent variable to be expressed dichotomously. With a Multivariate logistic regression analysis was used to evaluate independent predictors of poor outcomes. Accordingly, it was discovered that patients who had the following three characteristics were found to be more likely to have a poor outcome: pre-admit antimicrobial use (P=0.012, AOR 3.87(1.34-11.16)), patients with hospital stay above 10 days (P=0.029, AOR 3.49(1.13-10.72)), patients whose antimicrobial regimen changed (P=0.004, AOR 3.74(1.52-9.22.4)), patients with comorbidities (P=0.012, AOR 5.00(1.42-17.59)) (Table 10).

**Table 10.** Univariate and multivariate logistic regression analysis of factors associated with poor outcome among children with pneumonia who received antimicrobial therapy at Jimma Medical Center, Jimma, Ethiopia, from February 03 to June 03, 2022.

| Variables |  | Clinical outcome |  | COR (95% CI) | P-value | AOR (95% CI) | P-value |
| --- | --- | --- | --- | --- | --- | --- | --- |
|  |  | Good | Poor |  |  |  |  |
| Sex | Male | 57(63.3) | 33(36.7) | 1.222(0.603-2.476) | 0.577 |  |  |
|  | Female | 38(67.9) | 18(32.1) |  | 1 |  |  |
| Age category (in months) | <= 11 | 40(61.5) | 25(38.5) |  | 1 |  | 1 |
|  | 12-35 | 13(59.1) | 9(40.9) | 1.108(0.413-2.969) | 0.839 | 0.920(0.311-02.724) | 0.880 |
|  | 36-71 | 21(67.7) | 10(32.3) | 0.762(0.309-1.881) | 0.302 | 0.595(0.222-1.594) | 0.555 |
|  | >=72 | 24(85.7) | 4(14.3) | 0.267(0.083-0.860) | 0.027 | 0.337(0.097-1.175) | 0.088 |
| Weight (Kg) | 1-5.9 | 19(65.5) | 10(35.5) | 1.353(0.424-4.323) | 0.610 |  |  |
|  | 6-10.9 | 35(59.3) | 24(40.7) | 1.763(0.638-4.870) | 0.274 |  |  |
|  | 11-15.9 | 23(60.7) | 10(30.3) | 1.118(0.355-3.517) | 0.849 |  |  |
|  | >=16 | 18(72) | 7(28) | 1 | 1 |  |  |
| Residence | Rural | 85(65.9) | 44(34.1) | 0.739(0.263-2.076) | 0.567 |  |  |
|  | Urban | 10(58.8) | 7(41.2) | 1 | 1 |  |  |
| Admission type | Direct admission | 6(54.50) | 5(45.5) | 1 | 1 |  |  |
|  | Transferred from gov't institution | 79(65.8) | 41(34.2) | 0.623(0.179-2.164) | 0.456 |  |  |
|  | Transferred from private institution | 10(66.7) | 5(33.3) | 0.600(0.121-2.973) | 0.532 |  |  |
| Temperature | Febrile | 39(52.7) | 35(47.3) | 3.141(1.53-6.446) | 0.003 | 3.878(0.413-30.250) | 0.233 |
|  | afebrile | 56(77.7) | 16(22.3) |  | 1 |  | 1 |
| Respiratory Rate | Fast | 55(57.9) | 40(42.1) | 2.645(1.210-5.779) | 0.015 | 1.503(0.633-3.564) | 0.356 |
|  | Not fast | 40(78.4) | 11(21.6) |  | 1 |  | 1 |
| Oxygen saturation | <90% | 52(70.3) | 22(29.7) |  | 1 |  | 1 |
|  | >90% | 43(59.7) | 29(40.3) | 1.594(0.803-3.165) | 0.183 | 1.149(0.506-2.609) | 0.741 |
| Vaccination status | Full vaccinated | 47(57.3) | 35(42.7) |  | 1 |  |  |
|  | Partially vaccinated | 22(66.7) | 11(33.3) | 1.429(0.540-3.783) | 0.472 |  |  |
|  | Not vaccinated | 26(83.9) | 5(16.1) | 1.072(0.374-3.070) | 0.897 |  |  |
| Nutritional status | Normal | 66(70.2) | 28(29.8) |  | 1 |  |  |
|  | Malnourished | 29(55.8) | 23(44.2) | 1.869(0.925-3.770) | 0.081 | 3.247(0.785-13.245) | 0.104 |
| Type of pneumonia | Community acquired | 82(69.5) | 36(30.5) |  | 1 |  | 1 |
|  | Hospital-acquired | 6(54.5) | 5(45.5) | 1.898(0.544-6.624) | 0.315 | 1.745(0.050-11.054) | 0.830 |
|  | Aspiration-pneumonia | 7(41.2) | 10(58.8) | 3.254(1.147-9.228) | 0.027 | 2.051(1.045-94.525) | 0.713 |
| Severity of pneumonia | Severe pneumonia | 80(66.1) | 41(33.9) | 1.769(0.318-1.861) | 0.560 |  |  |
|  | Non-Severe pneumonia | 15(60) | 10(40) |  | 1 |  |  |
| Diagnostic method | CBC | 54(65.9) | 28(34.1) |  | 1 |  | 1 |
|  | Chest radiography | 3(100) | 0 | .000(0.000-) | 0.999 | 0.000(.000-) | 0.999 |
|  | CBC + Chest radiography | 32(66.7) | 16(33.3) | 0.964(0.454-2.050) | 0.925 | 1.087(0.431-2.742) | 0.860 |
|  | CBC + Chest radiography + Chest CT | 6(46.2) | 7(53.8) | 2.250(0.690-7.338) | 0.179 | 1.126(0.211-6.001) | 0.890 |
| Culture done | No | 60(60.6) | 39(39.4) |  | 1 |  | 1 |
|  | Yes | 35(74.5) | 12(25.5) | 1.896(0.878-4.093) | 0.103 | 1.264(0.529-3.017) | 0.598 |
| Co-morbidities | Yes | 64(57.7) | 47(27.8) | 5.691(1.881-17.22) | 0.002 | 5.004(1.423-17.594) | **0.012** |
|  | No | 17(4.7) | 21(55.3) |  | 1 |  | 1 |
| Pre-admit antibiotic use | Yes | 44(49.4) | 45(50.6) | 8.693(3.387-22.31) | 0.000 | 3.870(1.342-11.16) | **0.012** |
|  | No | 51(89.5) | 6(10.5) |  | 1 |  | 1 |
| Antimicrobial regimen changed | Yes | 29(45.5) | 35(54.7) | 4.978(2.387-10.38) | 0.000 | 3.746(1.522-9.222) | **0.004** |
|  | No | 66(80.5) | 16(19.5) |  | 1 |  | 1 |
| Duration of hospital stay | <=5 | 51(79.9) | 13(20.3) |  | 1 |  | 1 |
|  | 6-9 days | 27(69.2) | 12(30.8) | 2.7444(1.700-4.34) | 0.002 | 1.585(1.728-12.233) | 0.047 |
|  | >=10 days | 17(39.5) | 26(60.5) | 6.000(2.532-14.22) | 0.000 | 3.492(1.138-10.72) | **0.029** |

## 4. Discussion

Appropriate antimicrobial utilization is critical for antimicrobial resistance containment as well as good clinical and economic outcomes [14]. However, inappropriate uses of antimicrobials have a devastating effect on patients as well as the general population. The present study is one of the few from Ethiopia to assess practice on the use of antimicrobials to treat pneumonia in hospitalized children. Childhood pneumonia was selected because it is one of the most common infectious diseases in Jimma Medical Center.

The findings show that most children were treated without evidence of microbiological data. This result also showed the attending physician’s opinion on the use of microbiological studies, that more than three-quarters 15(78.9%) of the physicians hesitate to send culture and sensitivity tests. Recommendations from published guidelines for managing children with pneumonia recommend starting treatment with broad-spectrum antibiotics, but this should be accompanied by proper de- escalation based on culture results [20]. Published guidelines also recommend microbiological testing in the event of failure of oral or primary antimicrobial therapy in the treatment of children with pneumonia. This was the case for most study participants (83, 56.8%) in the current study. However, microbiological studies were performed on only 47(32.19%) patients. This result was lower than reports from Indonesia (43.5%) and Italy (98%) that had microbiological cultures obtained during hospitalization, respectively [21][22]. There are many possible causes for this. This may be a result of the poor attention of physicians to the microbiological data. As most physicians have stated, it is sufficient to treat children with pneumonia based on clinical information. Also, a loss of trust in achieving a positive culture result. This may also be because the laboratory may not be equipped with rapid diagnostic kits.

In the current study, all treatment approaches were found to be empirically initiated. Empiric antimicrobial therapy is generally categorized as appropriate (adequate) or inappropriate (inadequate) based on microbiological culture and susceptibility findings. An empirical treatment regimen is considered appropriate if the identified microorganism is sensitive to at least one of the prescribed antimicrobial agents [2]. Studies have shown that appropriate empiric antimicrobial therapy has been associated with decreased mortality in patients with many different types of infection. However, the lack of cultural and susceptibility data has devastating consequences for both patient and economic perspectives [23]. However, in this study, only five out of thirteen (38.4%) patients who received empiric antimicrobial regimens were seen to have culture results with appropriate coverage. This finding was higher than reports from Japan and Indonesia, which were (17%) and (15.7%) [24][21]. This might be due to the higher prevalence of pneumonia or the difficulty of differentiating between bacterial and viral infections in our study.

In this study, out of the 208 prescriptions, 129(88.4%) of the prescriptions were third generation cephalosporin. In addition, empirically, about 6.4% of patients received vancomycin. This result also showed most treating physician opinions in the preference of broad-spectrum antimicrobials over narrow-spectrum during initiating treatment for pneumonia 14(73.6%). This finding was in contrast with a report from Nepal in which ampicillin was the most common (70%) and broad-spectrum antibiotics such as linezolid, vancomycin, and meropenem were used in less than 1% of patients [25]. However, the result was in line with reports from Nekemte Referral Hospital (60%) that have shown the use of third-generation cephalosporins as the first-line treatment of children with pneumonia, respectively [26]. It was seen that several factors influenced antimicrobial selection. On the one hand, this was due to the lack of a standardized hospital-specific treatment guideline, which allowed prescribing physicians to use whatever antimicrobial agent they wanted. On the other hand, regardless of the type of pneumonia, the majority of study participants received broad-spectrum antibiotics like cephalosporin alone or in combination with other antibiotics during pre-hospital admission, which may influence physician antimicrobial selection. This may be due to the limited attitude of most treating physicians to the preference of a narrow spectrum; since patients already took antimicrobial prehospital, it’s enough reason to broaden the spectrum of antimicrobial regimens without proper consideration of patient details. On the other hand, it is due to the frequent stockouts of most of the antimicrobials in the hospital, and this led the treating physician to prescribe the available antimicrobial agents.

In the current study, 89 (60.9%) of the 146 patients received antibiotics after failing first-line antimicrobial therapy. Many studies have shown that prior antimicrobial drug exposure is associated with colonization and infection by resistant pathogens [27]. However, consideration of previous exposure before initiation of empiric antimicrobial treatment is not generally observed in this study, as evidenced by patients who were prescribed a similar antimicrobial receiving more than 72 hours of pre-admit therapy. This could be due to a lack of awareness of antimicrobial resistance and/or poor attention to this global and concerning issue. A study on inadequate antimicrobial treatment has shown that in-appropriateness of the empirical antimicrobial regimen was significantly associated with a higher mortality rate [28]. However, it was very difficult to evaluate the appropriateness of the rest of the patients’ antimicrobial therapy in this regard.

In the current study, many clinically stable and unstable patients (complicated cases) treated with broad-spectrum antimicrobial combinations completed a full-course initiated treatment. Antimicrobial de-escalation is a strategy for proper use of antimicrobial to balance empirical use and reduce the development of resistance. To that effect, antimicrobial use practices at JMC are increasing at an alarming rate and require coordinated immediate intervention. Therefore, unless continuous delivery of antimicrobial is an issue, proper strategies and balances should be maintained in the selection of empirical antimicrobial to minimize their overuse and/or unnecessary use.

The present study evaluated patient eligibility for IV to PO conversion using predefined criteria adopted from the SHEA guidelines [18]. Therefore, it was found that 32 patients were eligible for conversion. However, only three patients were transferred and the rest continued to receive IV antimicrobials. This may be due to a limited attitude to the benefits of switching from IV to PO by the treating physician. This was reflected in the answers to the question about the very limited practice of converting IV to PO. About 37% of treating physicians reported switching at discharge and IV administration antimicrobial in inpatients is a common practice (26.3%). Similar was demonstrated in a study in TASH, in which 82 (41.8%) of the treating physicians said that administration of IV antimicrobials for hospitalized patients is a usual practice in this hospital and conversion is only made at the time of discharge, and about 10 (5.1%) of the treating physicians also said IV is more effective than PO antimicrobials [29].

The current study also evaluated the guidelines used by treating physicians to prescribe antimicrobial for the management of children with pneumonia. Most physicians’ responses showed that they are using international guidelines and eBooks to treat children with pneumonia. No use of local guidelines was observed in this study. The reason behind this might be that the local standard treatment guidelines were not prepared taking into consideration tertiary care hospitals. It is also known that the guidelines were developed without actual local antibiogram data. This suggests that clinicians relied primarily on eBooks and international guidelines. Different guidelines and books have been published at different times, and their recommendations are often not the same [14]. This concept is also demonstrated in the guide to good prescribing, which was prepared by WHO [7]. This has diversified the use of antimicrobial in the treatment of patients diagnosed with similar pneumonia. The referenced eBooks and international guidelines are primarily based on national antimicrobial resistance patterns and are mostly for educational purposes. In addition, these eBooks and guidelines recommend having facility-specific guidelines based on the drug susceptibility of your own facility [2]. Several studies and published guidelines describe how the use of different strategies has been found helpful for more appropriate and cost-effective use of antimicrobial in hospitals. Such strategies include education, guidelines, and clinical pathways, antimicrobial order forms, de-escalation of therapy, intravenous-to-oral switch therapy, and dose optimization, common infection-based interventions such as community-acquired pneumonia [14][7][22].

In the current study, good or poor clinical outcome was the primary end point. Therefore, the observed finding is in-hospital mortality (11, 7.5 %). In addition, even though two or more combinations of antimicrobial were given for more than 10 days (43, 29.4%), patients were complicated (no improvement). This finding is higher than both results from the Nekemte Referral Hospital, Ethiopia, in which poor treatment was observed in 30.6% of pediatric patients [26] and from TASH, in which 7.7% of patients had poor treatment outcomes [29]. The findings of the present study suggest that the quality of care of patients with infectious diseases was minimal at JMC as compared to the previous study settings. Absence of a sufficient number of infectious diseases specialists, hospital specific antimicrobial treatment protocols, continuous supply of antimicrobials, and better microbiological laboratory services indicates poor quality care. This could also be due to a high proportion of comorbidities, such as cardiovascular disease and malnutrition, which can lead to a poor prognosis.

Identifying predictors for poor outcome was a critical strategy during infectious disease management in order to provide specialized care based on the number of risk factors associated with the patients. Hence, this study tried to assess the possible predictors of poor outcome in patients with pneumonia. Accordingly, around three predictors were identified, and these risk factors are in line with different studies done across the globe.

In this study, a history of antimicrobial use pre-admission (p = 0.012) was one of the predictors of poor clinical outcome. Patients who were exposed to antimicrobial prior to admission had 3.8 times worse outcomes than those who were not. This is because patients coming to JMC are referred from different lower health facilities and other hospitals according to the country’s referral structure or system. This can lead to colonization and subsequent infection by resistant bacteria [10]. As a result, clinicians caring for these patients should take their recent antimicrobial experience into account and balance empiric antimicrobial regimens for suspected infection until microbiological studies are completed.

In this study, initial antimicrobial regimen changed were associated with an increased incidence of poor outcomes compared to those who completed their entire course of therapy with the initial antimicrobial regimen. This may also be due to the patient’s antimicrobial changed being affected by resistant organisms to prescribed antimicrobials. Therefore, this factors that patient who had antimicrobial changed could have contributed to the poor outcomes as the difference was significant on multivariate logistic regression (p=0.004).

In the current study, another factor found to have a statistically significant association with poor outcomes was hospital stay duration. Hospital stay duration above ten (p = 0.029) were a negative predictor of clinical outcome in the management of pneumonia. Patients with increased hospital days above 10 days were about 3.4 times more likely to have a poor treatment outcome compared with patients who stay ≤ 5 days (AOR =3.49, 95% CI:2.2–143.1). Similar results were found in a TASH study, where patients who stayed 8 days were approximately 14.3 times more likely to have a poor treatment outcome than patients who stayed 3 days (AOR = 14.3, 95% CI: 1.35-151.1) [29].

### Limitation of study

Our study is not without limitations and should be interpreted with caution. To mention, this study was conducted at a single hospital, and the follow-up was also limited to the patient’s hospital stay.

## Conclusion

Empirical initiation of antimicrobial was seen completed without sufficient evidence of indication and microbiological and radiographic findings. More than one-fourth of the patients treated for pneumonia experienced poor outcomes, implicating the need for more attention during treatment. Generally, to improve appropriate antimicrobial use and patient clinical outcomes, the hospital needs a concerted effort from all concerned bodies and needs to establish a functional antimicrobial stewardship program as soon as possible

## Data Availability

All relevant data are within the manuscript and its supporting information files.

## Acknowledgements

We want to sincerely thank the study participants and their families/carers, who volunteered their time and all necessary information. We also would like to thank the data collectors and the pediatric wards staff of Jimma Medical Center for their cooperation and providing the requested information during the data collection period.

## Supporting information file

**S1 File.** Information Sheet, Written assent, Data collection format, and questionnaire for Study (Afan Oromo, Amharic, and English Version) (DOCX)

